# Evaluating the Reliability of a Custom GPT in Full-Text Screening of a Systematic Review

**DOI:** 10.1101/2025.02.04.25321655

**Authors:** Rachel C. Davis, Saskia S. List, Kendal G. Chappell, Ahmed Madar, Sigrun Henjum, Espen Heen

**Affiliations:** Department of Community Medicine and Global Health, University of Oslo, Oslo, Norway; Department of Nursing and Health Promotion, Oslo Metropolitan University, Oslo, Norway; Department of Nursing, Health and Laboratory Science, Ostfold University College, Oslo, Norway

**Keywords:** OpenAI, custom GPT, full text screening, large language models, systematic review, article filtration

## Abstract

Systematic reviewing is a time-consuming process that can be aided by artificial intelligence (AI). There are several AI options to assist with title/abstract screening, however options for full text screening (FTS) are limited. The objective of this study was to evaluate the reliability of a custom GPT (cGPT) for FTS.

A cGPT powered by OpenAI’s ChatGPT4o was trained and tested with a subset of articles assessed in duplicate by human reviewers. Outputs from the testing subset were coded to simulate cGPT as an autonomous and assistant reviewer. Cohen’s kappa was used to assess interrater agreement. The threshold for practical use was defined as a cGPT-human kappa score exceeding the lower bound of the confidence interval (CI) for the lowest human-human kappa score in inclusion/exclusion and exclusion reason decisions. cGPT as an assistant reviewer met this reliability threshold. With the Cohen’s kappa CI for human-human pairs ranging from 0.658 to 1.00 in the inclusion/exclusion decision, assistant cGPT and human kappa scores were encompassed in two of four pairings. In exclusion reason classification, the benchmark human-human kappa score CI range was 0.606 to 0.912. Assistant cGPT and human kappa scores were encompassed in one of four pairings. cGPT as an autonomous reviewer did not meet reliability thresholds.

cGPT as an assistant could speed up systematic reviewing in a sufficiently reliable way. More research is needed to establish standardized thresholds for practical use. While the current study dealt with physiological population parameters, cGPTs can assist in FTS of systematic reviews in any field.

**HIGHLIGHTS:**

- There are several AI options to assist in title/abstract screening in systematic reviewing, however, options for full text screening are limited.
- The reliability of a tailor-made AI model in the form of a custom GPT was explored in the role of an assistant to a human reviewer and as an autonomous reviewer.
- Interrater agreement was sufficient when the model operated in the role of assistant reviewer but not in the role of autonomous reviewer. Here the model misclassified two articles out of ten, whereas the human reviewers erred in approximately one out of ten articles.
- The study shows that it is possible to craft a custom GPT as a useful assistant in systematic reviews. cGPTs can be crafted to assist in reviews in any field.
- An automated setup for inputting articles and coding cGPT responses is needed to maximize the potential time-saving benefit.

## 1. BACKGROUND

Systematic reviews (SR) are critical for synthesizing evidence that should form the basis for up-to-date best practice and policy.^1^ Such a study of previously published work starts broad by identifying all potentially relevant articles. After a thorough screening of title/abstract and full text for inclusion, a narrow subset of studies remains. Typically, the screening and data-extraction stages happen in duplicate with two independent reviewers making judgements and reaching consensus when discrepancies occur. The process may translate to hundreds, if not thousands, of human work hours depending on the scope of the review.^2^ With the current explosion in artificial intelligence (AI) tools, there is now a high expectation for improved cost-efficiency.

Various AI tools are already present in SR software developed to assist in title/abstract and full text screening.^3^ Commonly, these tools provide predictions of article relevancy, however, do not provide determination of inclusion status. Nevertheless, their training parameters may serve as a guide when training conceptually novel AI models. Training material required to initiate relevancy predictions in these models ranges from 50 included and 50 excluded articles^4^ to a single article.^5^ The only AI tool currently available that will make determinations on inclusion or exclusion of an article is Nested Knowledge. It’s Robot Screener can be activated in the role of one of two reviewers when 40 excluded and 10 included articles have been categorized.^6^

It is generally agreed that the current capabilities of AI tools and models are not in a place to totally replace human reviewers for either title/abstract or full text screening.^7–10^ It has been reported that reliability of text mining AI tools is better for intervention studies than for complex health service topics.^7,9,11,12^

Performance metrics of AI in screening tasks are typically reported as sensitivity (recall), specificity, false negative rate, proportion missed, positive predictive value (precision), observed agreement, F1-scores, and time savings.^7,8,10–14^ Evaluation of inter-rater reliability between human and AI decisions can also be done using Cohen’s kappa statistical analysis. Kappa is a value between -1 and 1. A value of 0 indicates no agreement better than random chance between the raters. Perfect agreement or disagreement will yield +1.00 or -1.00, respectively. The benefit of kappa compared with the other metrics is that it adjusts for random agreement. ^15^ AI tools used in screening for reviews tend to have high specificity but lower (<90%) sensitivity.^11,14,16,17^ Performance metrics for binary outcomes expressed in this article are further defined in table 1.^12,14^

**Table 1.** Test performance metrics with mathematical definitions for binary outcomes.

| METRIC | ACRONYM | DEFINITION | EQUATION<sup>a</sup> |
| --- | --- | --- | --- |
| **Sensitivity (recall)** | SENS | the proportion of correctly included articles to all includable articles | $\frac{TP}{TP + FN}$ |
| **Specificity** | SPEC | the proportion of correctly excluded articles to all excludable articles | $\frac{TN}{TN + FP}$ |
| **False negative rate** | FNR | the proportion of incorrectly excluded articles to all includable articles | $\frac{FN}{TP + FN}$ |
| **Proportion missed** | PM | the proportion of incorrectly excluded articles to all excluded articles | $\frac{FN}{TN + FN}$ |
| **Positive predictive value (precision)** | PPV | the proportion of correctly included articles to all included articles | $\frac{TP}{TP + FP}$ |
| **Observed agreement (accuracy)** | OBS AG | the proportion of true positives and true negatives to all articles | $\frac{TP + TN}{TP + TN + FP + FN}$ |
| **F1** | -- | the harmonic mean of precision and recall, where 1 indicates perfect precision and recall and 0 indicates total failure of the model. | $\frac{2TP}{2TP + FP + FN}$ |

<sup>a</sup>True positive (TP): number of articles correctly included, True negative (TN): number of articles correctly excluded, False positive (FP): number of articles incorrectly included, False negative (FN): number of articles incorrectly excluded.

One promising AI model to aid in systematic reviewing is ChatGPT. ChatGPT is a large language model (LLM) with the ability to generate human-like text.^18^ A retrospective study of several different LLMs’ ability to identify inclusion criteria in title/abstract screening compared to human performance identified ChatGPT4o to be the model with the best combined sensitivity and specificity.^19^

In an early version of ChatGPT (ChatGPT3.5) Mehrabanian found a human-AI observed agreement of 76% when given a random subset of pre-screened title/abstracts.^20^ Alshami et al. evaluated the potential of ChatGPT3.5 to autonomously conduct SRs. In title/abstract screening, ChatGPT was able to identify unrelated articles with F1 scores ranging from 85% to 90%. Full text screening classification metrics were not reported. The major limitations of ChatGPT were identified in data extraction, limited recall, and incomplete responses.^8^ Purewal et al. used ChatGPT4o for screening and data extraction and reported a 75.6% sensitivity and 66.8% specificity in identification of inclusion status in full text screening. Out of eleven possible reasons, ChatGPT4o identified exclusion reason with an observed agreement of 40%, a result the authors reported may have been due to insufficient training.^13^ One must consider these limitations within the context of which version of ChatGPT was used. As of November 2023, it has been possible to create a custom GPT which tailors responses based on input instructions and uploaded files. These custom GPTs will not “forget” instructions in the same way as a non-custom ChatGPT model which begins each new conversation without background information on the specific question being asked.^21^ To the best of our knowledge, a scientific evaluation of a *custom* GPT’s performance in full text screening is yet to be published. Hence, the purpose of this study was to evaluate the reliability and time saving potential of a tailor-made custom GPT (cGPT) in full text screening within a SR. The work was motivated by the need to assess more than a thousand articles in full text. The outcome variables in the SR mainly appeared as intermediary outputs in the includable articles. This required the cGPT to identify defined concepts rather than just the primary studies’ main outcome(-s).^22^

## 2. METHODS

The articles used in this study were selected from an ongoing SR which began in 2023. Briefly, the SR aims to describe the global meta-mean and dispersion of its primary (population-level habitual 24-hour urine volume) and secondary (population-level habitual 24-hour creatinine excretion) outcomes, identified in primary studies of adult populations globally. Additionally, the association between several measurable background variables and these outcomes will be explored. ^22^ The SR’s corpus is screened in duplicate at the title/abstract and full text stages. The articles selected were published between 2020 and 2024 and consisted of a training and a testing set. Articles were coded into their inclusion **<u>status</u>** (‘included’ as 1, ‘excluded’ as 0) and **<u>exclusion</u>** **<u>reason</u>** (coded as 1-9). In the event of an included article, the reason was coded as 10. The list of exclusion reasons for coding cGPT responses can be viewed in table 2.

**Table 2.** Distribution of the different categories of exclusion from the human-human consensus.

| Exclusion reason |  | Explanation | Number of articles |
| --- | --- | --- | --- |
| 1 | Review<sup>a</sup> | Includes any type of review. | 10 |
| 2 | Population < 16 years<sup>b</sup> | Should not include individuals under 16 years old. | 0 |
| 3 | Non-free-living population | Includes hospitals, prisons, research facilities, and other confined settings. | 6 |
| 4 | Diseased population | That may distort representation of the general population.<sup>22</sup> | 13 |
| 5 | Condition affecting outcome | Such as manipulated diet or intravenous fluid administration. Does not include pregnancy/breastfeeding or manipulated 24-hour fluid intake if fluid intake is measured and reported. | 8 |
| 6 | N<30 | The includable population as described in 1-5 must have a minimum of 30 individuals. | 7 |
| 7 | Outcome data not collected | Studies which did not measure or collect all urine produced in 24 hours. | 41 |
| 8 | Outcome data not reported | 24h urine volume measured but unreported. | 84 |
| 9 | Awaiting classification | Not enough information within the article(s) of a study to make a determination. | 6 |
| 10 | Included article | None of the above apply. | 63 |
| **SUM** |  |  | **238** |

<sup>a</sup> Reviews were investigated for additional studies in full text before exclusion.
<sup>b</sup> Exclusion reason 2 was effectively filtered out during title/abstract screening and was not present at the full text stage.

The researcher responsible for uploading, recording, and interpreting responses from cGPT was the *cGPT operator*. cGPT’s performance was evaluated in two screening roles: (1) Autonomous reviewer, where cGPT’s analysis of inputted articles was coded without any interpretation by the operator; (2) Assistant reviewer, where the cGPT analysis was interpreted by the operator who decided to code the article in agreement with cGPT or not. cGPT’s ability as a reviewer in these roles was quantified and described as interrater agreement (Cohen’s kappa), observed agreement, classification performance metrics, and time savings. All screening took place within the Covidence environment, a platform for conducting screening and data extraction in reviews, between June and September 2024.

### 2.1. Aggregation of training and testing data

Beginning in the full text screening stage, four human reviewers (E. H., R. D., S. L., and K. G.) kept a log of article ID number, status, exclusion reason, study design, and field of study. All the articles were initially screened by E.H, whereas R. D., S. L., and K. G. together shared the work of the second screening. Once a total of 250 articles were classified and catalogued in duplicate by reviewers, a subset of 40 excluded and 10 included articles was selected as the training set. The articles purposefully encompassed all study designs, fields, and exclusion reasons possible to expose cGPT to the greatest variety of scenarios. The 200 test articles were augmented with an additional 38, representing all articles screened in duplicate by human reviewers at that point in time. The human-human consensus status of the testing set was 63 included and 175 excluded. Exclusion reasons for the testing set can be found in table 2.

### 2.2. Training

The first version of our cGPT was created on 23 August 2024 using the paid feature on OpenAI’s ChatGPT platform. The cGPT was powered by ChatGPT4o. Initially, cGPT received abridged versions of the protocol and relevant supplements with a short description included in its ‘Instructions’ text field in the ‘Configuration’ tab of the general custom GPT environment. cGPT provided incomplete and inconsistent responses when prompted to assess the training articles, and it was determined that a more structured prompt-response configuration would produce more accurate and complete responses. The complete configuration of cGPT can be found in Supplemental 1.

A flowchart was developed and uploaded to aid cGPT in navigating each article to a final decision (Supplemental 2). cGPT was instructed to ‘Navigate this article through the exclusion flowchart’. Its responses provided explanations for its reasoning for excluding or proceeding at each step of the flowchart. Errors in these responses were addressed by creating ‘patches’ in the ‘Instructions’ explicitly advising cGPT on how to proceed when faced with a certain scenario.

The team trialed differing levels of complexity with the flowchart. A persistent issue was cGPT’s reluctance to identify includable subpopulations within a study when following the flowchart, resulting in inappropriate exclusion of the study altogether. The simplest solution proved to be the best; cGPT’s primary prompt was changed to ‘Navigate each population in this article through the exclusion flowchart individually’.

### 2.3. Testing

Testing ran from 25 to 30 September 2024. The cGPT operator was blind to consensus status and exclusion reason decisions throughout testing. A new conversation with cGPT was opened and the 238 articles were uploaded one at a time using the final prompt, described previously. Conversations are separate windows that initially have no history of dialogue, like sending a new email instead of continuing in a thread. No feedback was provided to cGPT, and no other prompts were used for the duration of testing. The cGPT operator coded both cGPT’s raw evaluation of the articles as well as the operator’s interpretation of status and exclusion reason based on cGPT’s evaluation. The latter was supported by notes and tags left by the first human reviewer in the Covidence environment. Notes and tags could include pertinent information about the population, the context of the study, or the study outcomes. In this way, the roles of cGPT as an autonomous reviewer and cGPT as an assistant reviewer were coded in parallel. The testing set was assessed two separate times by cGPT so that kappa statistics could be calculated between cGPT test 1 and test 2. Test 1 was completed before beginning test 2.

Upon testing completion, the operator was unblinded and the decisions of the operator and cGPT were pooled from test 1 to identify the “best response”. This served as the testing results of cGPT in the assistant role. Best response was defined as selecting which response, either cGPT or the operator, agreed most closely with the consensus decision. The best response was meant to simulate what would happen in the live environment should a discrepancy occur between the operator and cGPT. In the case of such a discrepancy, the operator would select the decision believed to be most accurate and leave a note that the article could be classified in one of two ways. If the selected classification was in disagreement with the other human reviewer, consensus decision could be expedited by referring to the second possible classification described in the note.

### 2.4. Statistical Analysis

Acceptable performance of cGPT was determined by whether the interrater agreement (Cohen’s kappa point estimate) between cGPT and a human reviewer exceeded the lower bound of the confidence interval (CI) for the lowest human-human kappa score in both the status and exclusion reason decisions. The kappa score assumes no “true” classification in the interrater comparison. Therefore, we calculated additional classification performance metrics using the consensus as the “truth”, in accordance with the definitions in the introductory part of this article. These metrics were calculated from the binary variable “status”.

When performing kappa statistics for exclusion reason, both excluded articles with exclusion reasons coded 1 through 9 and included articles with reason coded 10 were included in analysis. Included articles were kept in the exclusion reason analysis due to the potential for discrepancy in the status decision between cGPT and the human reviewer.

R version 4.3.3 for Windows was used for all statistical calculation. Packages used in analysis included “psych” and “irr”. The observed agreement, kappa statistic confidence intervals, and classification performance metrics were calculated directly from equations in the R environment. Findings are reported in-text as point estimate (95% confidence interval).

## 3. RESULTS

### 3.1. Experiential observations

An important observation was that cGPT tended to “stumble”, a term coined by this team to describe the phenomenon whereby cGPT provided incomplete responses or failed to respond to the prompt altogether. When this occurred, the operator would reissue the prompt with the article attached. cGPT was observed to be more likely to stumble after it had stumbled a first time. It was also more likely to stumble the longer the conversation became, however, there did not appear to be a set length that would precipitate disrupted responses. Longer conversations also appeared to cause longer response times from cGPT. After observing these trends, it was determined that upon the first instance of cGPT stumbling, the most time-efficient response was to begin a new conversation. Test 1 required reinitiating the conversation four times to complete the testing set while test 2 required five.

cGPT was able to reliably divide populations within an article when prompted to do so before navigating populations through the exclusion flowchart. Engineering its prompt in this way allowed for simplification of the flowchart coupled with detailed ‘Instructions’. This combination produced responses that were complete and delivered in a prescribed format, with explanations for its decisions that allowed the cGPT operator to identify errors in its logic. In observational studies cGPT most often divided populations based on how the study reported its outcomes, and in intervention studies most often divided populations by intervention and control groups.

cGPT expressed a tendency to drift in its exclusion reason assessment, randomly favoring one reason over another. The most favored exclusion reasons were 3. (non-free-living population) and 5. (condition affecting urine volume). This random favoritism did not carry over when a new conversation was initiated.

cGPT at times would report logical fallacies within the boundaries of its responses. The three most common logical errors observed were: a population comprised of 30 or more individuals was classified as less than 30, resulting in an inappropriate exclusion reason 6; identifying that one of the outcomes was reported but excluding for reason 8 since the study did not report both of the outcomes; and inclusion based on reporting of urine analytes other than creatinine (calcium, iodine, etc.) in the absence of 24-hour urine volume reporting.

### 3.2. cGPT reproducibility

Between test 1 and test 2, the autonomous cGPT’s *status* decisions had an observed agreement of 0.777 (0.719, 0.829) and a Cohen’s kappa of 0.493 (0.368, 0.618). The autonomous cGPT’s *exclusion reason* decisions had an observed agreement of 0.622 (0.56, 0.684) and a Cohen’s kappa of 0.495 (0.429, 0.562).

### 3.3. Results of testing

The testing results are presented in the following way: classification performance metrics are presented using the binary variable “status” (table 3). This provides a gross overview of how cGPT compares to human reviewers when both are compared against the consensus decision (the “truth”).

**Table 3.**
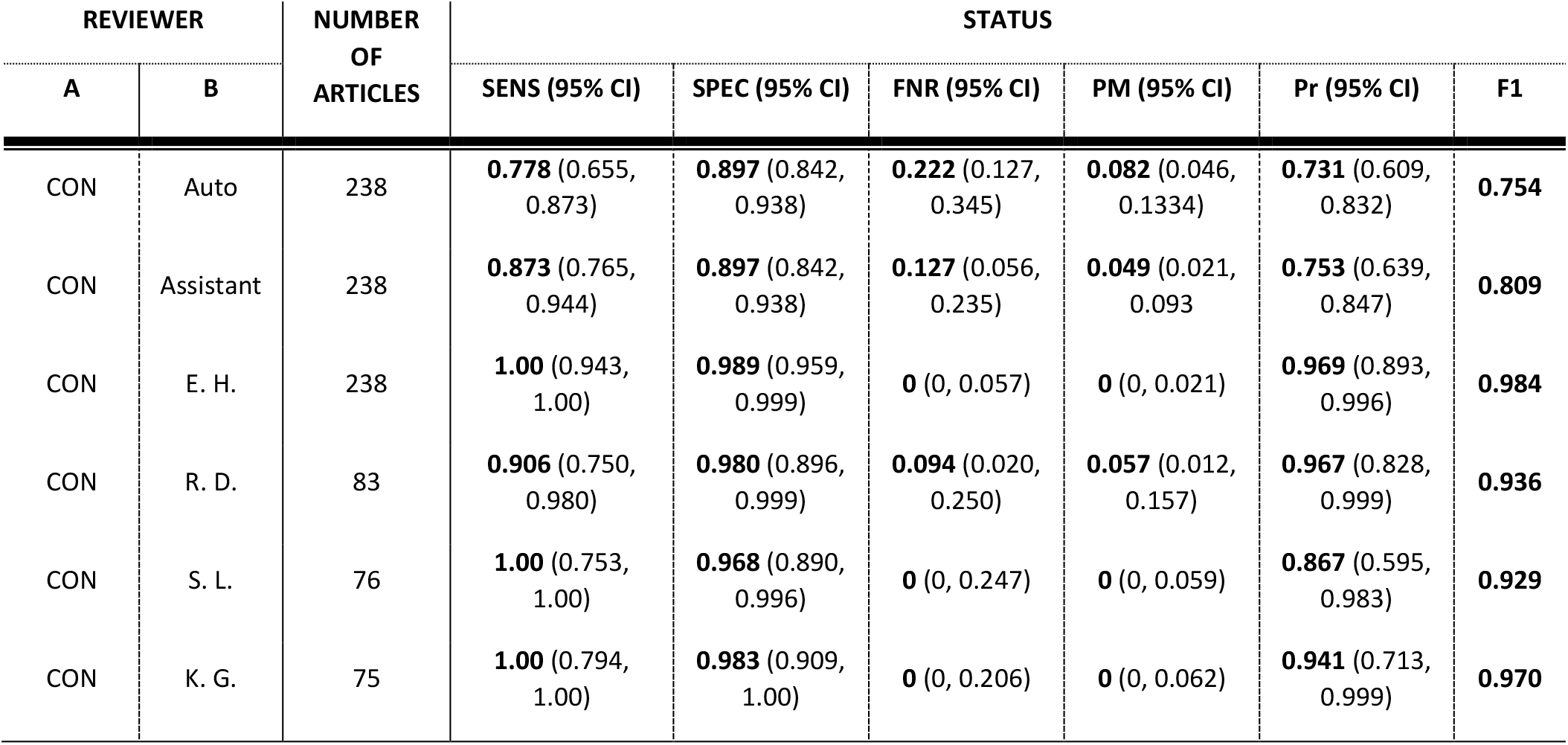
Classification performance metrics of <u>status</u> decisions. A: Consensus (CON) between human-human reviewers, cGPT as an autonomous reviewer (Auto), cGPT as an assistant reviewer (Assistant), sensitivity (SENS), specificity (SPEC), false negative rate (FNR), proportion missed (PM), precision (Pr).

Human-human kappa scores are presented, both with their status and exclusion reason decisions (table 4). These are the benchmarks to which cGPT in both its roles is compared. Table 4 also contains kappa scores (both status and exclusion reason decisions) of human reviewers paired with cGPT in the role of autonomous reviewer, followed by cGPT as an assistant reviewer.

**Table 4.** Observed agreement (OBS AG) and kappa scores of status and exclusion reason decisions between human-human pairings, autonomous cGPT-human pairings, and assistant cGPT-human pairings. Consensus (CON), cGPT as an autonomous reviewer (Auto), cGPT as an assistant reviewer (Assistant).

| REVIEWER |  | NUMBER<br>OF<br>ARTICLES | STATUS |  | EXCLUSION REASON |  |
| --- | --- | --- | --- | --- | --- | --- |
| A | B |  | OBS AG (95% CI) | COHEN'S KAPPA<br>(95% CI) | OBS AG (95% CI) | COHEN'S KAPPA<br>(95% CI) |
| E. H. | R. D. | 83 | **0.940** (0.865, 0.980) | **0.872** (0.658, 1.000) | **0.783** (0.694, 0.872) | **0.717** (0.611, 0.823) |
| E. H. | S. L. | 76 | **0.961** (0.889, 0.992) | **0.872** (0.647, 1.000) | **0.855** (0.776, 0.934) | **0.784** (0.656, 0.912) |
| E. H. | K. G. | 75 | **0.987** (0.928, 1.000) | **0.961** (0.735, 1.000) | **0.773** (0.679, 0.868) | **0.713** (0.606, 0.821) |
| Auto | CON | 238 | **0.866** (0.816, 0.906) | **0.661** (0.534, 0.788) | **0.651** (0.591, 0.712) | **0.533** (0.467, 0.599) |
| Auto | E. H. | 238 | **0.866** (0.816, 0.906) | **0.665** (0.539, 0.792) * | **0.655** (0.595, 0.716) | **0.532** (0.464, 0.600) |
| Auto | R. D. | 83 | **0.819** (0.720, 0.895) | **0.60** (0.385, 0.815) | **0.627** (0.522, 0.731) | **0.521** (0.417, 0.625) |
| Auto | S. L. | 76 | **0.882** (0.787, 0.944) | **0.668** (0.447, 0.889) * | **0.658** (0.551, 0.765) | **0.513** (0.387, 0.639) |
| Auto | K. G. | 75 | **0.853** (0.753, 0.924) | **0.59** (0.364, 0.816) | **0.60** (0.489, 0.711) | **0.465** (0.353, 0.577) |
| Assistant | CON | 238 | **0.891** (0.844, 0.927) | **0.733** (0.607, 0.859) | **0.719** (0.661, 0.776) | **0.632** (0.568, 0.696) |
| Assistant | E. H. | 238 | **0.882** (0.835, 0.920) | **0.715** (0.589, 0.841) * | **0.681** (0.621, 0.740) | **0.577** (0.511, 0.643) |
| Assistant | R. D. | 83 | **0.831** (0.733, 0.905) | **0.64** (0.425, 0.855) | **0.711** (0.613, 0.808) | **0.627** (0.523, 0.731) * |
| Assistant | S. L. | 76 | **0.882** (0.787, 0.944) | **0.623** (0.401, 0.845) | **0.658** (0.551, 0.765) | **0.519** (0.395, 0.643) |
| Assistant | K. G. | 75 | **0.88** (0.784, 0.944) | **0.678** (0.453, 0.903) * | **0.64** (0.531, 0.749) | **0.532** (0.422, 0.642) |

\*Satisfies reliability threshold.

#### 3.3.1. Classification performance metrics

The range of human reviewer status sensitivity was 0.906 (0.750, 0.980) to 1.00 (0.943, 1.00). cGPT as an autonomous reviewer produced sensitivity of 0.778 (0.655, 0.873) while cGPT as an assistant reviewer produced sensitivity of 0.873 (0.765, 0.944). For specificity, the human reviewer range was 0.968 (0.890, 0.996) to 0.989 (0.959, 0.999). cGPT as an autonomous reviewer produced a specificity of 0.897 (0.842, 0.938) as did cGPT as an assistant reviewer (0.897 [0.842, 0.938]), table 3.

#### 3.3.2. Human-human reviewer agreement

The point estimates of all human-human *status* kappa scores were between 0.872 (0.647, 1.00) and 0.961 (0.735, 1.00), hence the combined CI range was 0.647 to 1.00. The point estimates of all human-human reviewer *exclusion reason* kappa scores were 0.713 (0.606, 0.821) to 0.784 (0.656, 0.912), hence the combined CI range was 0.606 to 0.912, table 4 (top section). Human reviewer-consensus of observed agreement for *status* decisions ranged from 0.952 (0.881, 0.987) to 0.992 (0.970, 0.999) and observed agreement for *exclusion reason* decisions from 0.867 (0.790, 0.944) to 0.947 (0.897, 0.998).

#### 3.3.3. cGPT as an autonomous reviewer

The point estimate range of cGPT and individual human reviewer’s observed agreement for *status* decisions was 0.819 (0.720, 0.895) to 0.882 (0.787, 0.944), while Cohen’s kappa range was 0.59 (0.364, 0.816) to 0.668 (0.447, 0.889). Two Cohen’s kappa point estimates exceeded the lower bound of the human-human CI: 0.665 (0.539, 0.792) and 0.668 (0.447, 0.889). The point estimate range of cGPT and individual human reviewer’s observed agreement for *exclusion reason* decisions was 0.60 (0.489, 0.711) to 0.658 (0.551, 0.765), while Cohen’s kappa range was 0.465 (0.353, 0.577) to 0.532 (0.464, 0.600). No Cohen’s kappa point estimates exceeded the lower bound of the human-human CI table 4 (middle section).

#### 3.3.4. cGPT as an assistant reviewer

The point estimate range of the cGPT and individual human reviewer’s observed agreement for *status* decisions was 0.831 (0.733, 0.905) to 0.882 (0.835, 0.920), while Cohen’s kappa range was 0.623 (0.401, 0.845) to 0.715 (0.589, 0.841). Two Cohen’s kappa point estimates exceeded the lower bound of the human-human CI: 0.678 (0.453, 0.903) and 0.715 (0.589, 0.841). The point estimate range of cGPT and individual human reviewer’s observed agreement for *exclusion reason* decisions was 0.64 (0.531, 0.749) to 0.711 (0.613, 0.808), while Cohen’s kappa range was 0.519 (0.395, 0.643) to 0.627 (0.523, 0.731). One Cohen’s kappa point estimate exceeded the lower bound of the human-human CI: 0.627 (0.523, 0.731) table 4 (bottom section).

### 3.4. Time and workload savings

The time required to screen a fixed number of articles was determined by the total number of articles screened divided by the number of articles screened per hour. Time savings was calculated as the difference of time required to screen this set between human reviewers and cGPT. For cGPT, internet quality, type of article, number of populations within the article, and variability in generative speed of responses are unforeseen variables that impacted the number of articles screened per hour. For human reviewers, type of article (including whether the full text could be located), internet quality, and other environmental factors influenced how many articles were screened per hour. Due to this high variability, time savings are described as minimum and maximum. Out of six total observations, the minimum documented number of articles completed by a human reviewer in one hour was 9 while the maximum was 31. Out of four total observations, the minimum number of articles screened by cGPT was 32 and the maximum was 44. The 2020-2024 time-strata corpus of the present SR contains 1,057 full text articles for screening. Considering our minimum and maximum values, this translates to between 34.1 and 117.4 effective human hours and 24 to 33 cGPT hours to conduct a single screening on this time strata. The time savings then can be described as between 10.1 and 84.4 hours saved when using cGPT instead of a human reviewer. While the human reviewers can only keep their top screening speed for a few hours at a time, cGPT, by its nature, has only the duration limitations of the operator.

## 4. DISCUSSION

Kappa scores were our primary outcome of interest in determining which role cGPT was best suited for. The status kappa scores between cGPT as an autonomous reviewer and human status decisions were narrowly captured within the human-human kappa confidence interval in two out of four measurements, specifically autonomous cGPT paired with E. H. and S. L. Similarly, the status decision kappa scores of cGPT as an assistant and the human reviewers were within the confidence interval of human-human reviewer pairings in two out of four measurements, specifically assistant cGPT paired with E.H. and K.G.

Turning to the more complex exclusion reason decisions, none of the kappa scores between the autonomous cGPT reviewer and human reviewers were within the range of confidence intervals of human-human pairings, indicating that cGPT would not be appropriate to employ as an autonomous reviewer in this project for determining exclusion reasons. The exclusion reason kappa scores between the cGPT assisted reviewer and human reviewers was captured in one of four instances, namely the cGPT-assisted reviewer with R.D.

Consistent with what has been observed in another study of LLMs in SRs, cGPT did score below 90% sensitivity, with specificity outperforming sensitivity in both reviewer roles.^19^ Put plainly, cGPT is more likely to generate false negatives than false positives in status and exclusion reason decisions. The opposite can be observed for human reviewers, who in three out of four instances had higher sensitivity than specificity - meaning when in doubt, they included the article. Specificity was identical for both cGPT as an autonomous reviewer and assistant reviewer. Sensitivity was, however, higher in the assistant role, so the true strength of using cGPT as an assistant rather than as an autonomous reviewer lies in its ability to include more relevant articles, rather than to discard more irrelevant articles. This phenomenon is also reflected by a higher F1 score in cGPT as an assistant compared to cGPT as an autonomous reviewer.

Summarized, cGPT in the role of assistant reviewer provided sufficient interrater agreement scores to meet the minimum requirement for use in this project. cGPT’s lower sensitivity compared to other reviewers is partially mitigated by requiring screening to be done in duplicate, with one reviewer mandatorily being an unassisted human.

The study of LLMs like ChatGPT in full text screening is still largely uncharted territory with limited study comparability, pre-prints excluded. Indeed, the singular example of LLMs in full text screening located by the authors of this manuscript did not quantitatively report ChatGPT’s performance at this stage of screening but rather described experiences interfacing with the model.^8^ Further validation studies need to be conducted so results such as those presented here can be compared and a threshold for acceptable use established.

While Nested Knowledge is the only example of an AI tool explicitly designed for full text screening, its validation is reported almost exclusively with binary statistics (recall, precision, and F1), the exception being observed agreement.^13,23^ This does not allow for the nuance of exclusion reasons explored in our study, necessitating a more complex metric for interrater agreement. Furthermore, looking at cGPT’s roles as autonomous and assistant reviewer, observed agreement with the consensus status decision, used as the “truth”, was 86.6% and 89.1% respectively, compared to a human reviewer and consensus agreement range of 95.2% to 99.2%. This means that for the best human-consensus agreement, cGPT will miss one additional article for every ten articles screened. For exclusion reason, the cGPT and consensus pairing of observed agreement was 65.1% (autonomous cGPT) and 71.9% (assistant cGPT). The range of human reviewer and consensus exclusion reason agreement was 86.7% and 94.7%. While status agreement is quite similar across all pairings, the same cannot be said for exclusion reason agreement. To compare interrater agreement with ten alternative responses, a Cohen’s kappa is necessary. While there are no other studies to point to which have used Cohen’s kappa for this specific reason, the authors of this study felt it was the most appropriate metric to compare interrater agreement.

### 4.1. Limitations & future directions

There is risk for bias when using human decision as the “truth” by which to compare a screening tool. While this bias is partially mitigated by using the consensus decision (the power of two reviewers in agreement) as “truth”, there nevertheless remain borderline decisions that could be interpreted one of two ways. The kappa score captures this uncertainty within its metrics, while observed agreement does not.

cGPT was observed to randomly favor one exclusion reason over others. This bias is addressed in part by using cGPT as an assistant rather than as an autonomous reviewer, however since cGPT responses terminate when an exclusion reason is reached, the operator would be required to ask follow-up questions to discern the true exclusion reason. Since starting a new conversation with cGPT makes it “forget” this favoritism, a potential solution in the future may be to initiate a new conversation at fixed intervals.

cGPT at times would report logical fallacies within the boundaries of its responses. These logical fallacies likely contributed to the performance gap between cGPT as an autonomous reviewer and cGPT as an assistant reviewer. It is entirely possible that the novel ChatGPT models o1, o3-mini-high, and the coming GPT 5.0 will not make such mistakes.

Despite efforts to encourage reproducibility in its configuration, cGPT had a poorer kappa score with itself between test 1 and test 2 of the testing set than it did with consensus in either the assistant reviewer or autonomous reviewer role. Care must be taken by researchers who choose to employ a custom GPT for any purpose due to this inconsistency in its responses. It has been put forward that perhaps using a “majority vote” with 4 different LLMs evaluating articles together may produce improved sensitivity scores and could address the issue of inconsistent responses.^19^ Alternatively, it is possible that this issue may be abated by prompting cGPT with the same article five or more times and accepting the majority vote for classification. The present study was not streamlined to such a degree that requesting five iterations of analysis on a single article was feasible, nor were there resources enough to produce 4 different custom LLMs catered to the project. These may be opportunities to explore in the future.

cGPT’s testing as an assistant reviewer occurred after all 238 articles had been screened for a first time by human reviewers. The operator had the opportunity to read notes and tags left behind by the first human screener to confirm scoring decisions guided by cGPT. A team looking to employ a cGPT assisted reviewer would likely have the most advantageous results by also placing this individual in the role of second screener.

Since screening is done in duplicate, the potential time savings only covers half of the workload of full text screening. Even with the fastest examples of human reviewing compared to the slowest example of cGPT reviewing, cGPT still outperformed human reviewers’ screening speed. The question of whether the discrepancies generated by cGPT translate to a significant amount of time savings was not explored quantitatively here. Resolution of cGPT-generated discrepancies was first investigated by the cGPT operator. If the operator was in agreement with the human reviewer, resolution was reached by the operator without consulting the human reviewer. In practice, this saves time for the human reviewer who is not obliged to perform consensus on any but the most nuanced cases of disagreement. Even with the potential to save human work hours, time savings in vivo will depend on several factors including internet speed, availability and content of articles, and the unpredictability of cGPT generated responses as described throughout this article.

Since the outcomes of interest in our SR are mostly intermediary steps in the studies being extracted, the reporting of relevant outcomes can be less obvious than, for example, a review of drug outcome studies. If the SR’s outcome and the studies’ outcomes are aligned, cGPT may have performed even better with a kappa score closer to those of the human-human reviewer pairs.

The method used to develop the cGPT was designed to be one that researchers who do not have significant computer science background could reproduce or mimic. With a more technical background, a research team could possibly create cGPT as an application programming interface (API) that would pull in articles from a cloud server, thus negating the need for an operator to manually upload one article at a time into the cGPT conversation.^19,24^ The purpose of the present study was both to evaluate the legitimacy of cGPT as a full text screener as well as describe the time saving potential of the method used. While its reliability is evaluated in detail here, further exploration into optimizing cGPT’s timesaving potential is needed.

The present SR is a considerably large study with data from 1990 to 2024 and thousands of articles requiring full text screening. It was feasible to take several hundred articles for the use of testing and training an AI model that can then be employed in the remaining articles. Many SRs will not have screening needs as demanding as those presented here, so there is likely a minimum threshold for size of SRs where the benefits of using a custom GPT outweigh the initial time investment to train and test it.

Using LLMs like ChatGPT in this way does not come without cost. There are environmental impacts, financial costs, social consequences, and the potential for a disruption in what is prioritized in this type of research.^25,26^ While the production of energy required to power the servers that support LLMs is becoming greener, it is unclear whether the costs outweigh the benefits from a larger systems perspective.

## 5. CONCLUSION

cGPT performed best when employed as a reviewer’s assistant and met reliability thresholds, although not consistently. More research is needed to establish standardized reliability guidelines for practical use of custom GPTs. There is evidence of time saving potential to utilizing cGPT. A more sophisticated system for inputting articles and coding cGPT’s responses is needed to maximize the potential time-saving benefits. Investigating cGPT’s role as either an assistant or autonomous reviewer in a less complex SR may provide improved reliability and time saving potential which outpace the current study.

## Supporting information

Supplemental 1: Instructions

Supplemental 2: Flowchart

## CONTRIBUTIONS

All members of the team contributed to conceptualization, data curation, reviewing, and editing. Rachel Davis was responsible for methodology, software, formal analysis, investigation, resources, visualization, original draft writing, and project administration. Espen Heen was instrumental in methodology, visualization, validation, project administration, editing, and supervision.

## ACKNOWLEDGEMENT

Haakon Meyer provided useful comments to the manuscript.

## USE OF ARTIFICIAL INTELLIGENCE TOOLS

A custom GPT (Pro-version), powered by ChatGPT4o from OpenAI, was used as described throughout this article, but was not used in the generation of text , tables, or in any other content of the article.

## ETHICS STATEMENT

Not applicable.

## CONFLICTS OF INTEREST

The authors have no conflicts of interest to disclose.

## FUNDING STATEMENT

This study did not receive any funding.

## DATA AVAILABILITY STATEMENT

All data produced in the present study are available upon reasonable request to the authors.

## Notes

### Competing Interest Statement

The authors have declared no competing interest.

### Summary of Updates

Manuscript adapted for journal publication. Assessment of appropriateness of use in the current project modified from cGPT-consensus kappa score surpassing the lower bound of the CI of the lowest scoring human-human kappa, to a cGPT-human reviewer kappa score surpassing the lower bound of the CI of the lowest scoring human-human kappa.

## REFERENCES

1. Gopalakrishnan S, Ganeshkumar P. Systematic reviews and meta-analysis: Understanding the best evidence in primary healthcare. J Fam Med Prim Care. 2013;2(1):9–14. doi:10.4103/2249-4863.109934

2. Borah R, Brown AW, Capers PL, Kaiser KA. Analysis of the time and workers needed to conduct systematic reviews of medical interventions using data from the PROSPERO registry. BMJ Open. 2017;7(2):e012545. doi:10.1136/bmjopen-2016-012545

3. Ge L, Agrawal R, Singer M, et al. Leveraging artificial intelligence to enhance systematic reviews in health research: advanced tools and challenges. Syst Rev. 2024;13(1):269–7. doi:10.1186/s13643-024-02682-2

4. Ouzzani M, Hammady H, Fedorowicz Z, Elmagarmid A. Rayyan-a web and mobile app for systematic reviews. Syst Rev. 2016;5(1):210–210. doi:10.1186/s13643-016-0384-4

5. van de Schoot R, de Bruin J, Schram R, et al. An open source machine learning framework for efficient and transparent systematic reviews. Nat Mach Intell. 2021;3(2):125–133. doi:10.1038/s42256-020-00287-7

6. Robot Screener – Nested Knowledge. Accessed January 22, 2025. https://about.nested-knowledge.com/docs/robot-screener/

7. Giummarra MJ, Lau G, Gabbe BJ. Evaluation of text mining to reduce screening workload for injury-focused systematic reviews. Inj Prev. 2020;26(1):55–60. doi:10.1136/injuryprev-2019-043247

8. Alshami A, Elsayed M, Ali E, Eltoukhy AEE, Zayed T. Harnessing the Power of ChatGPT for Automating Systematic Review Process: Methodology, Case Study, Limitations, and Future Directions. Syst Basel. 2023;11(7):351. doi:10.3390/systems11070351

9. Paynter R, Bañez LL, Erinoff E, Lege-Matsuura J, Potter S. Commentary on EPC methods: an exploration of the use of text-mining software in systematic reviews. J Clin Epidemiol. 2017;84:33–36. doi:10.1016/j.jclinepi.2016.11.019

10. van Dijk SHB, Brusse-Keizer MGJ, Bucsán CC, van der Palen J, Doggen CJM, Lenferink A. Artificial intelligence in systematic reviews: promising when appropriately used. BMJ Open. 2023;13(7):e072254–e072254. doi:10.1136/bmjopen-2023-072254

11. Gates A, Johnson C, Hartling L. Technology-assisted title and abstract screening for systematic reviews: A retrospective evaluation of the Abstrackr machine learning tool. Syst Rev. 2018;7(1):45–45. doi:10.1186/s13643-018-0707-8

12. Valizadeh A, Moassefi M, Nakhostin-Ansari A, et al. Abstract screening using the automated tool Rayyan: results of effectiveness in three diagnostic test accuracy systematic reviews. BMC Med Res Methodol. 2022;22(1):160–160. doi:10.1186/s12874-022-01631-8

13. Purewal A, Fautsch K, Klasova J, Hussain N, D’Souza RS. Human versus artificial intelligence: evaluating ChatGPT’s performance in conducting published systematic reviews with meta-analysis in chronic pain research. Reg Anesth Pain Med. Published online 2025:rapm-2024-106358. doi:10.1136/rapm-2024-106358

14. dos Reis AHS, de Oliveira ALM, Fritsch C, Zouch J, Ferreira P, Polese JC. Usefulness of machine learning softwares to screen titles of systematic reviews: a methodological study. Syst Rev. 2023;12(1):68–68. doi:10.1186/s13643-023-02231-3

15. Landis JR, Koch GG. The Measurement of Observer Agreement for Categorical Data. Biometrics. 1977;33(1):159–174. doi:10.2307/2529310

16. Rathbone J, Hoffmann T, Glasziou P. Faster title and abstract screening? Evaluating Abstrackr, a semi-automated online screening program for systematic reviewers. Syst Rev. 2015;4(1):80–80. doi:10.1186/s13643-015-0067-6

17. Pham B, Jovanovic J, Bagheri E, et al. Text mining to support abstract screening for knowledge syntheses: a semi-automated workflow. Syst Rev. 2021;10(1):1–156. doi:10.1186/s13643-021-01700-x

18. Roumeliotis KI, Tselikas ND. ChatGPT and Open-AI Models: A Preliminary Review. Future Internet. 2023;15(6):192. doi:10.3390/fi15060192

19. Li M, Sun J, Tan X. Evaluating the effectiveness of large language models in abstract screening: a comparative analysis. Syst Rev. 2024;13(1):219–17. doi:10.1186/s13643-024-02609-x

20. Mehrabanian M. AI-based article screening. Br Dent J. 2023;235(12):914–915. doi:10.1038/s41415-023-6692-x

21. Introducing GPTs. Accessed January 22, 2025. https://openai.com/index/introducing-gpts/

22. Heen E, Madar A, Meyer H, et al. 24-hour urine volume and 24-hour urine creatinine excretion in general adult populations, a systematic review and meta-analysis. Published online January 17, 2024. doi:10.17605/OSF.IO/GQZA3

23. Sauca M. AI for Screening: How is it used, and how can we tell how accurate it is? January 23, 2025. Accessed January 23, 2025. https://about.nested-knowledge.com/2024/08/02/ai-for-screening-how-is-it-used-and-how-can-we-tell-how-accurate-it-is/

24. Guo E, Gupta M, Deng J, Park YJ, Paget M, Naugler C. Automated Paper Screening for Clinical Reviews Using Large Language Models: Data Analysis Study. J Med Internet Res. 2024;26(2):e48996. doi:10.2196/48996

25. Bender EM, Gebru T, McMillan-Major A, Shmitchell S. On the Dangers of Stochastic Parrots: Can Language Models Be Too Big? In: New York, NY, USA: ACM; 2021:610–623. doi:10.1145/3442188.3445922

26. Rillig MC, Ågerstrand M, Bi M, Gould KA, Sauerland U. Risks and Benefits of Large Language Models for the Environment. Env Sci Technol. 2023;57(9):3464–3466. doi:10.1021/acs.est.3c01106

