## Supplemental 2: Flowchart for "Evaluating the Reliability of a Custom GPT in Full-Text Screening of a Systematic Review"

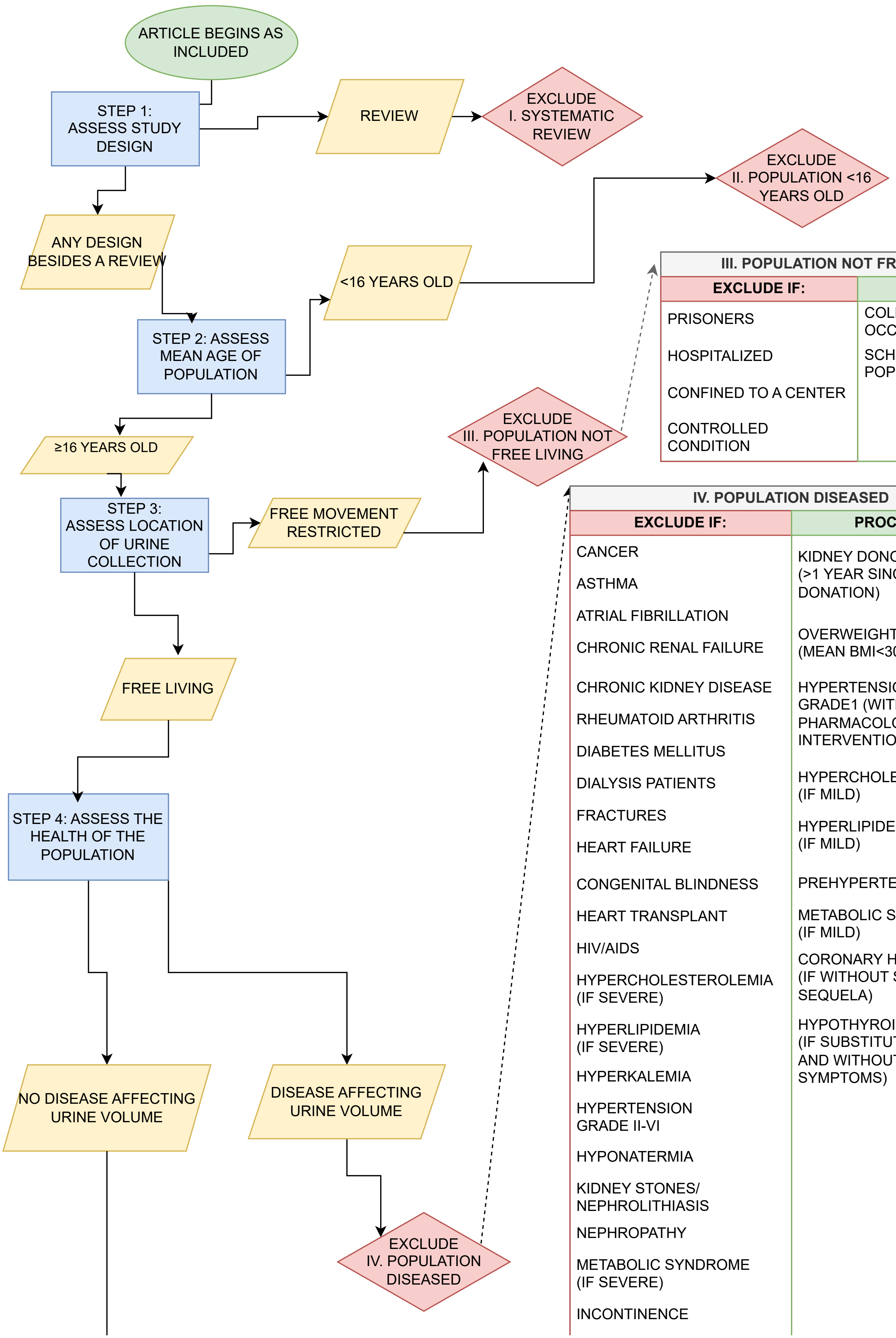

**III. POPULATION NOT FREE LIVING**

| EXCLUDE IF: | PROCEED IF: |
|---|---|
| PRISONERS | COLLECTION OCCURRED AT HOME |
| HOSPITALIZED | SCHOOL-BASED POPULATION |
| CONFINED TO A CENTER | |
| CONTROLLED CONDITION | |

**IV. POPULATION DISEASED**

| EXCLUDE IF: | PROCEED IF: |
|---|---|
| CANCER | KIDNEY DONOR (>1 YEAR SINCE DONATION) |
| ASTHMA | OVERWEIGHT (MEAN BMI<30) |
| ATRIAL FIBRILLATION | HYPERTENSION GRADE1 (WITHOUT PHARMACOLOGICAL INTERVENTION) |
| CHRONIC RENAL FAILURE | HYPERCHOLESTEROLEMIA (IF MILD) |
| CHRONIC KIDNEY DISEASE | HYPERLIPIDEMIA (IF MILD) |
| RHEUMATOID ARTHRITIS | PREHYPERTENSION |
| DIABETES MELLITUS | METABOLIC SYNDROME (IF MILD) |
| DIALYSIS PATIENTS | CORONARY HEART DISEASE (IF WITHOUT SYMPTOMS OR SEQUELA) |
| FRACTURES | HYPOTHYROIDISM (IF SUBSTITUTED AND WITHOUT SYMPTOMS) |
| HEART FAILURE | |
| CONGENITAL BLINDNESS | |
| HEART TRANSPLANT | |
| HIV/AIDS | |
| HYPERCHOLESTEROLEMIA (IF SEVERE) | |
| HYPERLIPIDEMIA (IF SEVERE) | |
| HYPERKALEMIA | |
| HYPERTENSION GRADE II-VI | |
| HYPONATERMIA | |
| KIDNEY STONES/ NEPHROLITHIASIS | |
| NEPHROPATHY | |
| METABOLIC SYNDROME (IF SEVERE) | |
| INCONTINENCE | |

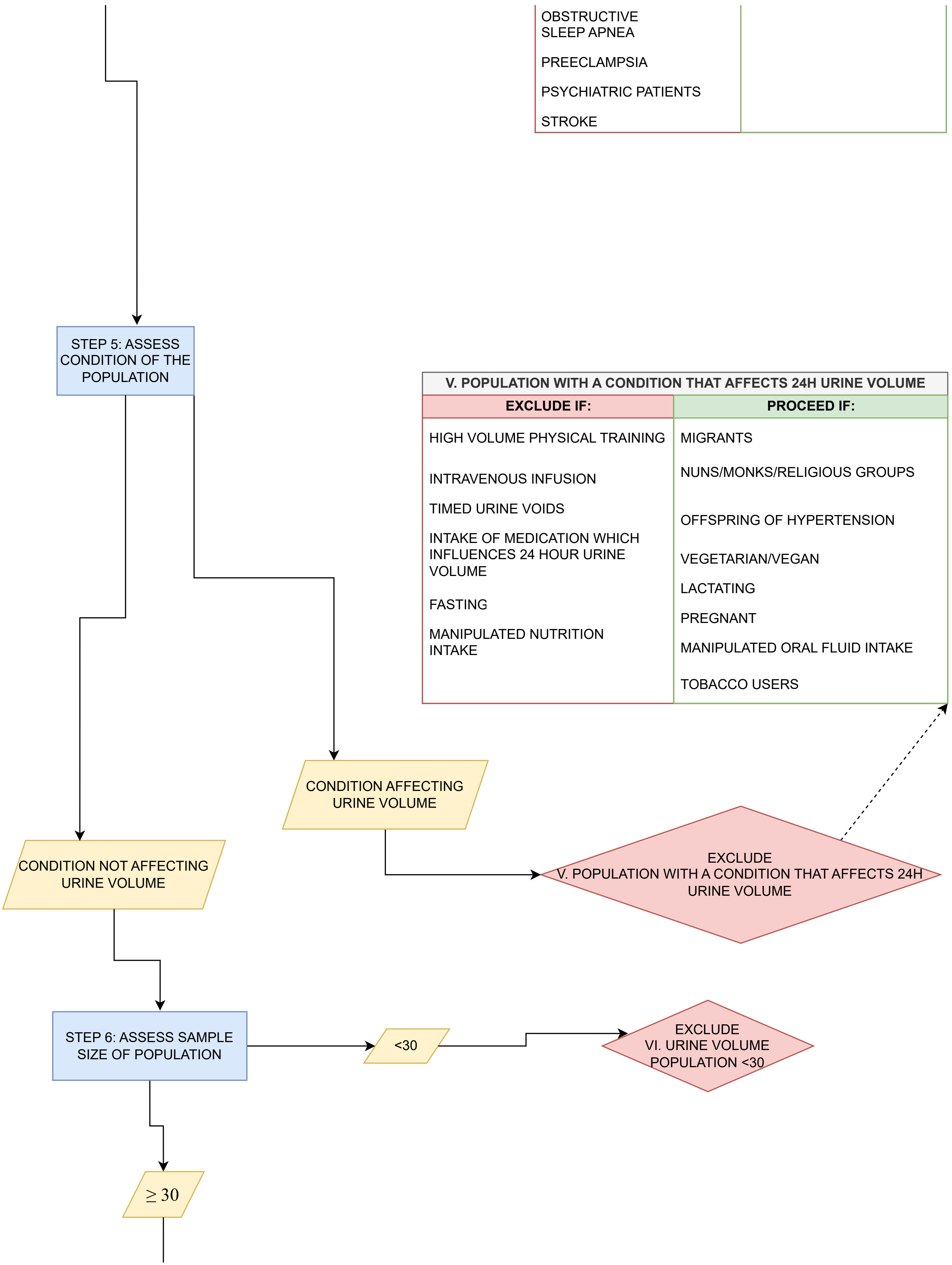

| |
|---|
| OBSTRUCTIVE SLEEP APNEA |
| PREECLAMPSIA |
| PSYCHIATRIC PATIENTS |
| STROKE |

**V. POPULATION WITH A CONDITION THAT AFFECTS 24H URINE VOLUME**

| EXCLUDE IF: | PROCEED IF: |
|---|---|
| HIGH VOLUME PHYSICAL TRAINING | MIGRANTS |
| INTRAVENOUS INFUSION | NUNS/MONKS/RELIGIOUS GROUPS |
| TIMED URINE VOIDS | OFFSPRING OF HYPERTENSION |
| INTAKE OF MEDICATION WHICH INFLUENCES 24 HOUR URINE VOLUME | VEGETARIAN/VEGAN |
| FASTING | LACTATING |
| MANIPULATED NUTRITION INTAKE | PREGNANT |
| | MANIPULATED ORAL FLUID INTAKE |
| | TOBACCO USERS |

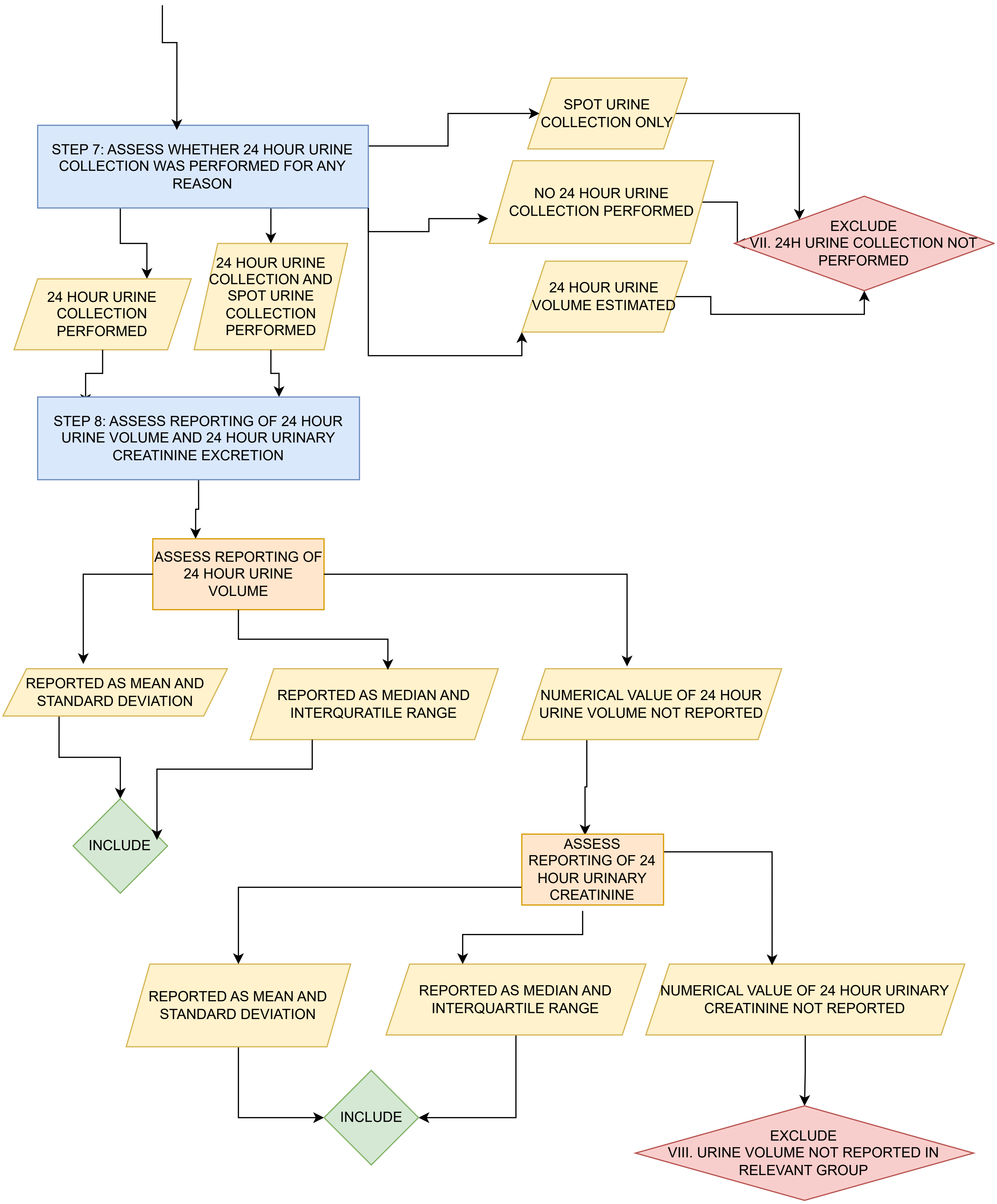
STEP 7: ASSESS WHETHER 24 HOUR URINE COLLECTION WAS PERFORMED FOR ANY REASON
SPOT URINE COLLECTION ONLY
NO 24 HOUR URINE COLLECTION PERFORMED
24 HOUR URINE VOLUME ESTIMATED
EXCLUDE
VII. 24H URINE COLLECTION NOT PERFORMED
24 HOUR URINE COLLECTION PERFORMED
24 HOUR URINE COLLECTION AND SPOT URINE COLLECTION PERFORMED
STEP 8: ASSESS REPORTING OF 24 HOUR URINE VOLUME AND 24 HOUR URINARY CREATINE EXCRETION
ASSESS REPORTING OF 24 HOUR URINE VOLUME
REPORTED AS MEAN AND STANDARD DEVIATION
REPORTED AS MEDIAN AND INTERQURATILE RANGE
NUMERICAL VALUE OF 24 HOUR URINE VOLUME NOT REPORTED
INCLUDE
ASSESS REPORTING OF 24 HOUR URINARY CREATININE
REPORTED AS MEAN AND STANDARD DEVIATION
REPORTED AS MEDIAN AND INTERQUARTILE RANGE
NUMERICAL VALUE OF 24 HOUR URINARY CREATININE NOT REPORTED
INCLUDE
EXCLUDE
VIII. URINE VOLUME NOT REPORTED IN RELEVANT GROUP
